# Evaluation of the Impact of a 90-day Mortality Prediction Model Integrated in the EHR for Advanced Care Planning^*^

**DOI:** 10.1101/2024.10.08.24315123

**Authors:** Lorenzo A. Rossi, Laura Roberts, Marinel Olivares, Amis Christian, Stefanie Mooney, Denise Morse, Finly Zachariah

## Abstract

Medical centers have begun to integrate AI models for mortality prediction in their electronic health records (EHR) systems to support advance care planning.^1^ There is evidence that advance care planning can reduce suffering and unwanted, costly treatment near the end of life.^2^

To better target patients at a risk of death with advance directive (AD) capture and goals of care (GOC) conversations at our comprehensive cancer center, we developed and integrated a 90-day mortality prediction model in the EHR. The model has been live since February, 2021. Performance in production has matched very closely pre-production pilots, achieving an 85% AUROC (AI Showcase Stage I). We implemented 4 clinical decision support (CDS) tools driven by the model to promote goal concordant care in inpatient and outpatient encounters (AI Showcase Stage II).^3^

In this document, we analyze the impact of one of the aforementioned CDS applications, implemented as an interruptive alert for the clinical social workers (CSWs) to facilitate advance directive (AD) completion in the inpatient setting. We also present preliminary results on volumes of GOC conversations prompted by one of the model-driven alerts. Our analysis is based on 12 months of data, collected between March 1, 2021 and February 28, 2022.

The challenges for our assessment included inconsistent EHR documentation practices, coexistence with (traditional) non-AI driven processes tasked for similar goals and many external factors that can affect target outcomes. We believe such challenges to be rather common across healthcare organizations integrating AI in their workflows.

## Methods

Our analysis focused patients admitted without an AD. The AD interruptive alert went live on 2.16.2021, targeting adult inpatients without an AD in place (roughly 2/3 of the admissions). CSWs have historically relied upon a mix of criteria to prioritize patients to approach for AD completion, ranging from seriousness of the condition to lack of clarity in healthcare agents, because of insufficient capacity to approach every admitted patient. Such criteria remained in place even after alert go-live. The process of AD completion initiated in the hospital is often completed after discharge *e*.*g*. to allow patients and families more time to process the documents. On a filed AD there is no indication if a CSW facilitated the process. The aforementioned factors lead to uncertainties on the number of AD completions that (1) can be associated with the hospital stay and, (2) can be credited to the model driven alert.

Based on feedback from domain experts, we assumed all the ADs completed between admission and up to 14 days after patient discharge to be associated with the hospital stay (either because of the model or other processes). The assumption was further validated after inspecting the distribution of the counts of ADs completed with post-discharge tolerance windows ranging from 0 to 28 days. As implied, some of the ADs filed after the firing of the alert may be actually due to pre-existing (non-AI based) processes. To increase our confidence on the actual use of the alert, we compared rates of the AD completions across acknowledgement reasons selected by the CSWs when prompted with the alert. Since an alert may fire more than once for a patient encounter, we focused on the last acknowledgment reason given (Fig. 1a). Then for simplicity, we credited every AD filed after an alert to the model.

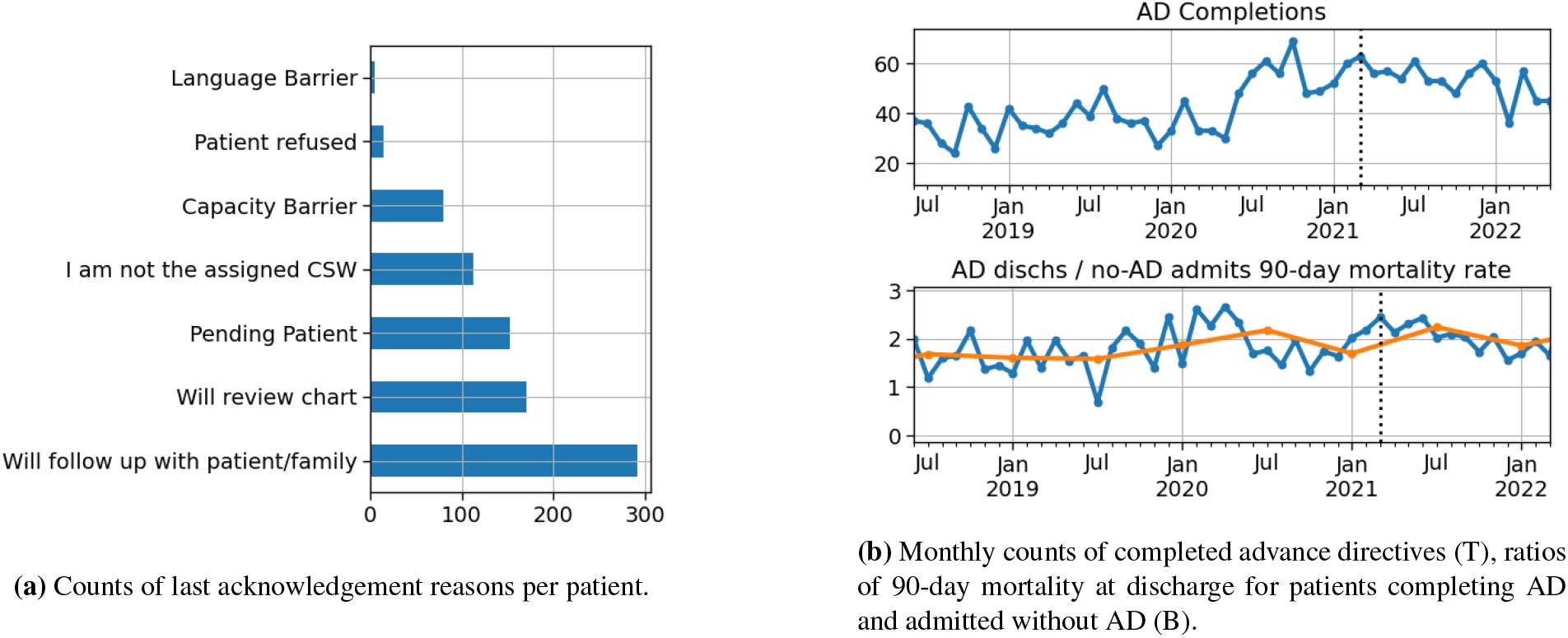

We computed the monthly counts of completed ADs and ratios of 90-day mortality rates for patients who completed AD to the 90-day mortality rates for all the patients without AD (both evaluated at discharge) (Fig. 1b). The latter is to measure performance of the AD process in targeting patients at high risk of death: the metric would be approximately equal to one if the patients solicited for AD completions were chose randomly.

## Results

We observed the highest rate of AD completion per alert when the last acknowledgment reason was “I will follow up with patient/family” (27%, *p* = .008) and a relatively lower AD completion rate when the last acknowledgement reason was “I’m not the assigned CSW” (15%, *p* = .048).

We computed counts of patients with completed ADs, for which we evaluated metrics including length of stay (LOS), in-hospital mortality rates for patients with AD completed and the aforementioned ratio of 90-day mortality rate at discharge (Tab. 1).

**Table 1.** Advance Directives completed after firing of the alert vs. ones completed without alert involved.

|  | Dates | Encounters | Patients | ADs | AD LOS<br>Median (SD) | Days to AD (SD) | AD Inpt.<br>Mortality % | 90-day<br>Mortality Ratio |
| --- | --- | --- | --- | --- | --- | --- | --- | --- |
| **AI Alert** | 3.01.21 - 2.28.22 | 788 | 643 | 166 | 12 (25.7) | 6 (19) | 14.5 | **4.55** |
| **No AI Alert** | 3.01.21 - 2.28.22 | 4410 | 3046 | 477 | 8 (14.2) | 4 (8.4) | 3.1 | 1.13 |
| **All** | 3.01.21 - 2.28.22 | 5198 | 3399 | 644 | 9 (18.4) | 6 (15.2) | 6.1 | **2.02** |
| **Year before AI** | 3.01.20 - 2.28.21 | 5097 | 3258 | 604 | 11 (21.2) | 6 (15.4) | 7.0 | 1.86 |

We also examined a GOC interruptive alert. The alert was used to order 169 supportive care consults (with an associated 73% Precision) for GOC in its first year and resulted in at least 67 documented GOC discussions.

## Discussion

The results in Tab. 1 and Fig. 1b and additional analyses not included here suggest that up to half of the AD completions attributed to the alert could have been facilitated via non-AI driven approaches. Our study showed that the model has been used by the clinical social workers as an additional tool to facilitate advance directive completion for inpatients at higher risk of death, facilitating up to the 25.8% of the ADs in a year, The tool showed a significant superior ability to target patients nearing end-of-life thanks to the accuracy of the underlying model. We have been making adjustments (i.e. increasing decision threshold of the AD alert) to increase use and impact of the alert. A model-driven GOC alert also resulted in documentation of GOC for a subset of patients that may otherwise not have been prioritized for such discussions.

## Data Availability

The data associated with the current study is not publicly available as the data are not legally certified as being de-identified.

